# Relationship between Serum Leptin, Lipid Metabolism, HbA1c, and Renal Function in Individuals with Type 2 Diabetes Mellitus and Obesity and in Individuals with Type 2 Diabetes Mellitus without Obesity

**DOI:** 10.1101/2023.02.20.23286131

**Authors:** A. A. Akinjinmi, A.A. Amballi, Abdulrahman Abdulbasit Opeyemi, Aderinto Nicholas, Adeniyi Akinbobola Ayokunle, I. A. Akindele, AbdulBasit Opeyemi Muili

## Abstract

**Background:** Type 2 diabetes mellitus (T2DM) is a chronic metabolic disorder characterised by hyperglycaemia, impaired lipid metabolism, and insulin resistance. In addition to the traditional risk factors, recent studies have suggested that adipose tissue-derived hormones such as Leptin play a crucial role in the pathogenesis of T2DM and its related complications. However, the exact mechanism underlying the relationship between serum leptin levels and these comorbidities remains unclear.

**Methods and Materials:** The study included 222 individuals, with 74 having T2DM and obesity, and 74 having T2DM without obesity and 74 control group. Serum levels of leptin, total cholesterol, triglycerides, high-density lipoprotein (HDL), low-density lipoprotein (LDL), and HbA1c were measured, and the glomerular filtration rate (GFR) was used as a measure of renal function.

**Results:** Serum leptin levels were significantly higher in individuals with T2DM and obesity compared to those without obesity (p<0.05). Additionally, total cholesterol, triglycerides, and LDL were significantly higher in individuals with T2DM and obesity compared to those without obesity (p<0.05). Serum leptin levels were positively correlated with total cholesterol, triglycerides, and LDL (p<0.05) and negatively correlated with HDL (p<0.01). The GFR was not significant when both groups were compared.

**Conclusion:** Obesity has a negative impact on lipid profile and glycemic control in type 2 diabetes patients and highlights the significance of considering the effect of obesity on the relationship between leptin and lipid profile in diabetic patients. These findings suggest that targeting obesity and leptin levels may be an effective strategy for improving lipid profile and glycemic control in type 2 diabetes.

## BACKGROUND

Type 2 diabetes mellitus (T2DM) is a chronic metabolic disorder characterised by hyperglycaemia, impaired lipid metabolism, and insulin resistance (1). The prevalence of T2DM has been steadily increasing worldwide, and it is a major public health concern due to its association with various comorbidities, including cardiovascular disease (CVD) and chronic kidney disease (CKD) (2). In addition to the traditional risk factors, recent studies have suggested that adipose tissue-derived hormones such as Leptin play a crucial role in the pathogenesis of T2DM and its related complications (3, 4).

Leptin is a hormone secreted by adipose tissue that regulates energy balance and appetite by suppressing hunger and promoting energy expenditure (5). It has been reported that serum leptin levels are elevated in individuals with obesity and T2DM (6, 7). Furthermore, several studies have suggested that hyperleptinaemia is associated with impaired lipid metabolism and renal dysfunction in T2DM (8, 9). However, the exact mechanism underlying the relationship between serum leptin levels and these comorbidities remains unclear.

This study aims to investigate the relationship between serum leptin, lipid metabolism, HbA1c, and renal function in individuals with T2DM and obesity and individuals with T2DM without obesity. We hypothesise that serum leptin levels are associated with impaired lipid metabolism and renal function in individuals with T2DM and that this association may be influenced by the presence of obesity.

## METHODOLOGY

### Study Patients

Two hundred and twenty-two (222) individuals of both sexes aged 18–65 years (seventy-four (74) obese individuals (aged 49.8 ± 10.5 years) and seventy-four (74) non-obese individuals (aged 47.9 ± 9.6), both with type 2 diabetes mellitus, who consecutively visited the diabetes clinic of the Olabisi Onabanjo University Teaching Hospital, Sagamu, Ogun State, Nigeria, and seventy-four (74) screened individuals (aged 48.3 ± 11.2 years) that do not have diabetes, were also recruited as control individuals from the patients at the general outpatient clinic of the same health institution for three months, T2DM was defined based on the history of patients taking oral hypoglycemic drugs or according to the classification of the American Diabetes Association as showing a fasting plasma glucose concentration greater than 126 mg/dL (1). Individuals with diabetes were treated with oral hypoglycemic agents (metformin, n = 30, glibenclamide, n = 11). No patients received insulin therapy. None of the individuals suffered from significant renal, hepatic, or cardiovascular diseases. Diabetes lasted between 1 and 6 years (mean: 2.50± 1.44 years). Patients did not consume alcohol or perform heavy exercises for at least one week before the study.

The non-diabetic control group included 74 middle-aged and elderly non-obese people aged 48.3± 11.2 years who had received an annual health check-up. The following criteria were used to select the non-diabetic control individuals: 1) No diabetes in their first-degree relatives, 2) Fasting plasma glucose level less than 110 mg/dL, 3) haemoglobin A1c concentration less than 5.5%. Non-diabetic subjects with endocrine disease, significant renal or hepatic diseases, or those receiving medications that control glucose metabolism, hypertension, or hyperlipidemia were excluded from the study. Non-obesity was defined according to WHO criteria (BMI <30 kg/m2) (2).

### Anthropometric Evaluation

Indicators of anthropometry such as height, weight, hip circumference, and waist circumference were measured while subjects were standing and dressed comfortably without shoes. Body mass index (BMI) was determined based on height and weight measurements in kilograms and centimetres, respectively. The hip circumference was measured at the largest standing horizontal circumference of the buttocks, and the waist circumference was measured at the lowest standing horizontal circumference between the lower margin of the rib cage and the iliac crest. The waist-to-hip ratio (WHR) was also determined by dividing hip circumference by waist circumference, which is the ratio of waist circumference to hip circumference in cm (3). Well-trained nutritionists measured these variables. The medical officer thoroughly examined the participants to determine their current state of health.

### Analytical Methods

A total of ten millilitres (10 mL) of blood were taken from the test subjects and the control group, respectively, after an overnight fast of 10–12 hours. The blood samples were dispensed in fluoride-oxalate bottles for glucose, EDTA bottles for glycosylated haemoglobin, and serum-separating tubes for Leptin, urea, creatinine, and lipid profile.

The serum was separated immediately, aliquoted into plain bottles, and stored at -20 oC until ready for Analysis. The blood samples were analysed for fasting plasma glucose using the glucose oxidase enzymatic method, leptin (ELISA kit), and lipid profile using the Randox Test

Kit, the Urease method for urea, and the Jaffe method for creatinine. All analytical assays carried out followed the manufacturer’s instructions.

### Biochemical Analysis

After the subjects fasted for 12 hours, 10 mL of blood was taken through a vein. The blood samples were dispensed in fluoride oxalate bottles for glucose, EDTA bottles for glycosylated haemoglobin, and serum-separating tubes for Leptin, urea, creatinine, and lipid profile. The plasma and serum were separated, aliquoted into plain bottles, and stored at -20 oC immediately until ready for Analysis. The blood samples were analysed for fasting plasma glucose using the glucose oxidase enzymatic method, leptin (ELISA kit), and lipid profile using the Randox Test Kit, the Urease method for urea, and the Jaffe method for creatinine. Glycated haemoglobin (HbA1c) was measured immunoturbidimetrically using a microparticle agglutination inhibition method. All analytical assays carried out followed the manufacturer’s instructions. The low-density lipoprotein (LDL-C) cholesterol level was calculated using the Friedewald formula (LDL cholesterol = total CholesterolCholesterol - HDL cholesterol - 1/5 triglycerides) in subjects with serum triglyceride concentrations less than 400 mg/mL. Serum leptin concentration was measured by enzyme-linked immunosorbent assay (ELISA) with a commercially available human leptin ELISA kit (Bio Vendor Laboratory Medicine, Inc., GmbH) using specific human leptin antibodies. The intra- and inter-assay coefficients of variation were less than 5% for Leptin. Before the assay, quality controls and all sera were diluted ⅓ times with a diluting buffer.

### Ethical Approval

The Olabisi Onabanjo University Teaching Hospital’s Health Research Ethics Committee evaluated and approved the study, and all individuals provided written informed consent following a description of the method.

## RESULT

### Biophysical Profiles

Table 1.1 shows the mean values and standard deviations of the biophysical measurements of diabetic non-obese subjects and diabetic obese subjects. There was no statistically significant difference (p> 0.05) between the mean age of diabetic non-obese (47.9± 9.6) compared with diabetic obese (49.8 ± 10.5). There was a statistically significant difference (p < 0.05) between the body mass index of diabetic non-obese (23.4 ± 1.6) compared with diabetic obese (33.7 ± 3.7). There was also a statistically significant difference (p < 0.05) between the weight (kg) of diabetic non-obese (66.7 ± 8.2) compared with diabetic obese (90.1± 12.7). The waist-hip ratio of the diabetic non-obese subjects (0.85 ± 0.05) compared with diabetic obese (0.87 ± 0.09) showed a statistically significant difference (p <0.05).

**Table 1.1.** Biophysical measurements of DM non-obese and DM obese (Mean ± SD)

| **PARAMETER**<br>**S** | **DIABETIC**<br>**NON-OBESE**<br>**Mean ± SD** | **DM OBESE**<br>**Mean ± SD** | **z-test** | **p-VALUE** |
| --- | --- | --- | --- | --- |
| **Age (Yrs)** | 47.9± 9.6 | 49.8 ± 10.5 | -0.951 | 0.524 |
| **Weight (Kg)** | 66.7 ± 8.2 | 90.1 ± 12.7 | -13.171 | 0.000* |
| **BMI (kg/m<sup>2</sup>)** | 23.4 ± 1.6 | 33.7 ± 3.7 | -21.008 | 0.000* |
| **WHR** | 0.85 ± 0.05 | 0.87 ± 0.09 | -4.311 | 0.015* |

Level of significance $P < 0.05$

Table 1.3 shows the mean values and standard deviations of the Biophysical measurements of Diabetic obese subjects and controls. There was no statistically significant difference (p> 0.05) between the mean age of diabetic obese (49.8 ± 10.5) compared with controls (48.3 ± 11.2). There was a statistically significant difference (p < 0.05) between the body mass index of diabetic obese (33.7 ± 3.7) compared with controls (23.3 ± 1.6). There was also a statistically significant difference (p < 0.05) between the weight (kg) of diabetic obese (90.1 ± 12.7) compared with controls (65.5 ± 8.2). The waist-hip ratio of the diabetic obese subjects (0.87 ± 0.09) to controls (0.85 ± 0.06) showed a statistically significant difference (p >0.05).

Table 1.2 shows the mean values and standard deviations of the Biophysical measurements of diabetic non-obese subjects and control. There were no statistically significant differences (p> 0.05) between the mean age of diabetic non-obese (47.9± 9.6) compared with controls (48.3 ± 11.2) (p > 0.05), body mass index of diabetic non-obese (23.4 ± 1.6) compared with controls (23.3 ± 1.6)(p > 0.05), weight (kg) of diabetic non-obese (66.7 ± 8.2) compared with controls (65.5 ± 8.2) (p > 0.05). The waist-hip ratio of the diabetic non-obese subjects (0.85 ± 0.05) compared with controls (0.85±0.06) also showed no statistically significant difference (p < 0.05).

**Table 1.2.**
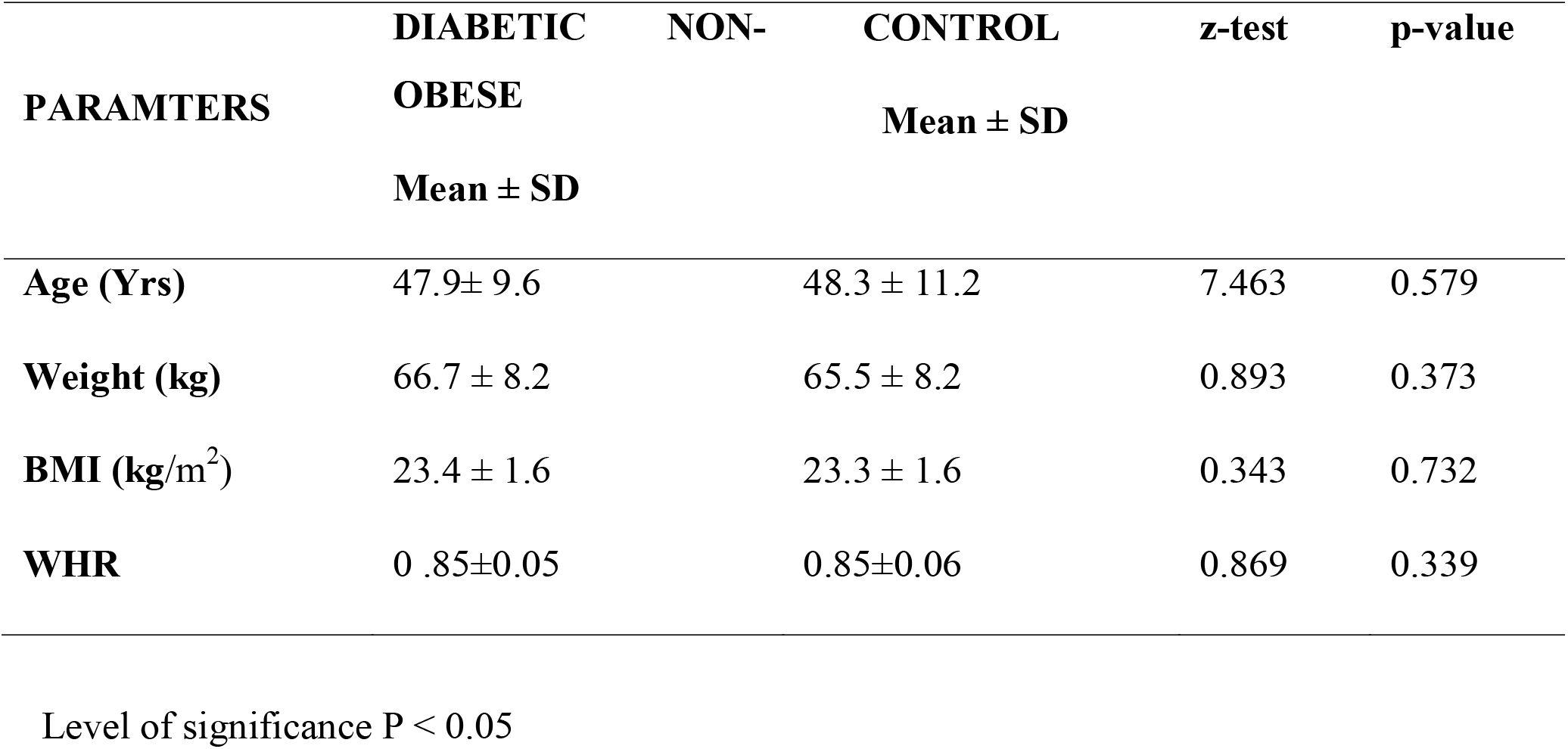
Biophysical measurements of DM non-obese, DM obese and control (Mean ± SD)

**Table 1.3.** Biophysical measurements of DM Obese and control (Mean ± SD)

| PARAMTERS | DIABETIC OBESE | CONTROL | z-test | p-value |
| --- | --- | --- | --- | --- |
| | Mean ± SD | Mean ± SD | | |
| **Age** | 49.8 ± 10.5 | 48.3 ± 11.2 | 3.329 | 0.233 |
| **Weight (kg)** | 90.1 ± 12.7 | 65.5 ± 8.2 | 13.673 | 0.000* |
| **BMI** | 33.7 ± 3.7 | 23.3 ± 1.6 | 21.193 | 0.000* |
| **WHR** | 0.87±0.09 | 0.85±0.06 | 2.705 | 0.000* |

Level of significance $P < 0.05$

Table 1.4 shows the mean values and standard deviations of test and control subjects’ biophysical measurements. There were no statistically significant differences (p > 0.05) between the mean ages of diabetic non-obese subjects (47.9± 9.6) and diabetic obese subjects (49.8 ± 10.5) compared with controls (48.3 ± 11.2). There were statistically significant differences (p < 0.05) between the body mass index of diabetic non-obese subjects (23.4 ± 1.6) and diabetic obese subjects (33.7 ± 3.7) compared with controls (23.3 ± 1.6). The waist-hip ratio of the diabetic non-obese subjects (0.85 ± 0.05) and diabetic obese subjects (0.87 ± 0.09) compared with controls (0.85 ± 0.06) showed a statistically significant difference (p < 0.05).

**Table 1.4.** Biophysical measurements of DM non-obese, DM Obese and control (Mean ± SD) using ANOVA.

| **PARAMETER**<br>**S** | **DIABETIC**<br>**NON-OBESE**<br>**Mean ± SD** | **DM OBESE**<br>**Mean ± SD** | **Control**<br>**Mean ± SD** | **F.RATIO** | **P-VALUE** |
| --- | --- | --- | --- | --- | --- |
| **Age** | 47.9± 9.6 | 49.8 ± 10.5 | 48.3 ± 11.2 | 54.151 | 0.472 |
| **Weight (Kg)** | 66.7 ± 8.2 | 90.1 ± 12.7 | 65.5 ± 8.2 | 28.925 | 0.000* |
| **BMI** | 23.4 ± 1.6 | 33.7 ± 3.7 | 23.3 ± 1.6 | 6.157 | 0.000* |
| **WHR** | 0.85 ± 0.05 | 0.87 ± 0.09 | 0.85 ±0.06 | 4.311 | 0.015* |

Level of significance $P < 0.05$

### Biochemical Profiles

Table 2.1 shows the mean values and comparisons of the biochemical parameters of diabetic non-obese subjects and diabetic obese subjects. There was no statistical difference (p>0.05) between the mean Triglyceride (TG) levels of the diabetic non-obese subjects (102.0 ± 41.7) compared with diabetic obese (105.8 ± 48.6). There were statistically significant differences (p<0.05) between the mean values of HbA1c of diabetic non-obese subjects (6.7 ± 1.1) compared with diabetic obese subjects (8.0 ±1.9), Leptin of diabetic non-obese subjects(6.1 ± 5.1) compared with diabetic obese subjects (8.1 ±6.5) (p<0.05); total CholesterolCholesterol (TCHOL) of diabetic non-obese subjects (192.1 ± 33.6) compared with diabetic obese subjects (221.9 ± 52.0); high-density lipoprotein (HDL) of diabetic non-obese subjects (54.3 ± 23.7) compared with diabetic obese subjects (45.1 ± 17.0); low-density lipoprotein (LDL) of diabetic non-obese subjects (103.0 ± 31.2) and diabetic obese (155.3 ± 56.0)

**Table 2.1.** Biochemical parameters of DM non-obese and DM obese subjects (Mean ± SD)

| PARAMETERS | DIABETIC<br>NON-OBESE<br>Mean ± SD | DIABETIC<br>OBESE<br>Mean ± SD | z-test | p. value |
| --- | --- | --- | --- | --- |
| HbA1c (%) | $6.7 \pm 1.1$ | $8.0 \pm 1.9$ | -4.974. | .000* |
| LEPTIN(1.18-3.82ng/ml) | $6.1 \pm 5.1$ | $8.1 \pm 6.5$ | -2.056 | .042* |
| TCHOL(130-230mg/dl) | $192.1 \pm 33.6$ | $221.9 \pm 52.0$ | -4.143 | .000* |
| TRIG (30-170mg/dl) | $102.0 \pm 41.7$ | $105.8 \pm 48.6$ | -.518 | .605 |
| HDL(30-70mg/dl) | $54.3 \pm 23.7$ | $45.1 \pm 17.0$ | 2.710 | .008* |
| LDL(100-159mg/dl) | $103.0 \pm 31.2$ | $155.3 \pm 56.0$ | -7.020 | .000* |
| UREA(0-50mg/dl) | $31.3 \pm 9.8$ | $33.4 \pm 10.9$ | 1.724 | .087 |
| CREAT(0.5-1.2mg/dl) | .85 ± 2.3 | .87 ± 11.9 | -1.072 | .285 |
| FPG | 125.8 ± 45.6 | 128.4 ± 44.0 | -352 | .725 |
| GFRml/min | 111.1 ± 25.00 | 110.5 ± 39.2 | -352 | .725 |

Level of significance $p < 0.05$

Table 2.2 shows the mean values and comparisons of the biochemical parameters of diabetic non-obese subjects and controls. There was no statistically significant difference between the mean Leptin of diabetic non-obese subjects (6.1 ± 5.1) compared with controls (5.3 ±5.9), high-density lipoprotein (HDL) of diabetic non-obese subjects (54.3 ± 23.7) compared with controls (51.7 ± 16.2). There were statistically significant differences (p<0.05) between the mean values of HbA1c of diabetic non-obese subjects (6.7 ± 1.1) compared with control (4.9 ±.73); Total Cholesterol (TCHOL)of diabetic non-obese subjects (192.1 ± 33.6) compared with controls (204.9 ± 43.9); low-density lipoprotein (LDL) of diabetic non-obese subjects (103.0 ± 31.2) compared with control subjects (133.5 ± 45.4); FPG of diabetic non-obese subjects (125.8 ± 45.6) compared with control subjects (88.7± 4.6) (p<0.05)

**Table 2.2.**
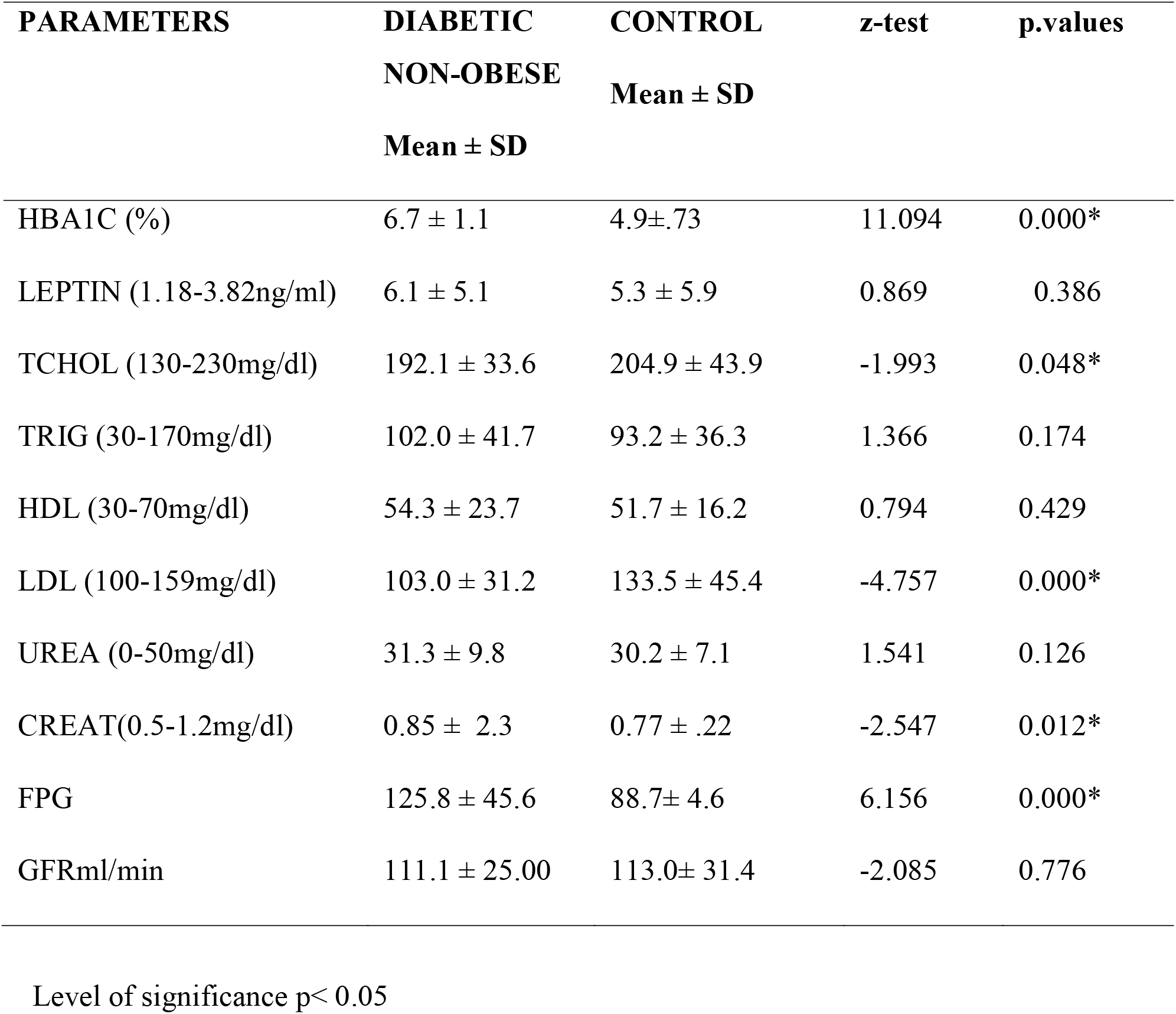
Biochemical parameters of DM non-obese and Control subjects (Mean ± SD)

| **PARAMETERS** | **DIABETIC**<br>**NON-OBESE**<br>**Mean ± SD** | **CONTROL**<br>**Mean ± SD** | **z-test** | **p.values** |
| --- | --- | --- | --- | --- |
| HBA1C (%) | 6.7 ± 1.1 | 4.9±.73 | 11.094 | 0.000* |
| LEPTIN (1.18-3.82ng/ml) | 6.1 ± 5.1 | 5.3 ± 5.9 | 0.869 | 0.386 |
| TCHOL (130-230mg/dl) | 192.1 ± 33.6 | 204.9 ± 43.9 | -1.993 | 0.048* |
| TRIG (30-170mg/dl) | 102.0 ± 41.7 | 93.2 ± 36.3 | 1.366 | 0.174 |
| HDL (30-70mg/dl) | 54.3 ± 23.7 | 51.7 ± 16.2 | 0.794 | 0.429 |
| LDL (100-159mg/dl) | 103.0 ± 31.2 | 133.5 ± 45.4 | -4.757 | 0.000* |
| UREA (0-50mg/dl) | 31.3 ± 9.8 | 30.2 ± 7.1 | 1.541 | 0.126 |
| CREAT(0.5-1.2mg/dl) | 0.85 ± 2.3 | 0.77 ± .22 | -2.547 | 0.012* |
| FPG | 125.8 ± 45.6 | 88.7± 4.6 | 6.156 | 0.000* |
| GFRml/min | 111.1 ± 25.00 | 113.0± 31.4 | -2.085 | 0.776 |

Level of significance $p < 0.05$

Table 2.3 shows the mean values and comparisons of the biochemical parameters of obese diabetic subjects and controls. There was no statistically significant difference between the mean Triglyceride of diabetic obese subjects (105.8 ± 48.6)and controls (93.2 ± 36.3). There were statistically significant differences (p<0.05) between the mean values of HbA1c of diabetic obese subjects (8.0 ±1.9) and control (4.9 ±0.73); Leptin of diabetic obese subjects (8.1 ± 6.5)and Controls (5.3 ± 5.9); Total Cholesterol (TCHOL)of diabetic obese subjects (221.9 ± 52.0) and Controls subjects (204.9 ± 43.9); high-density lipoprotein (HDL) of diabetic obese subjects(45.1 ± 17.0) and Controls (51.7 ± 16.2); low-density lipoprotein (LDL) of diabetic obese subjects(155.3 ± 56.0) and Control Subjects (133.5 ± 45.4)); FPG of diabetic obese subjects(128.4 ± 44.0) and Control Subjects (88.7 ± 4.6).

**Table 2.3.**
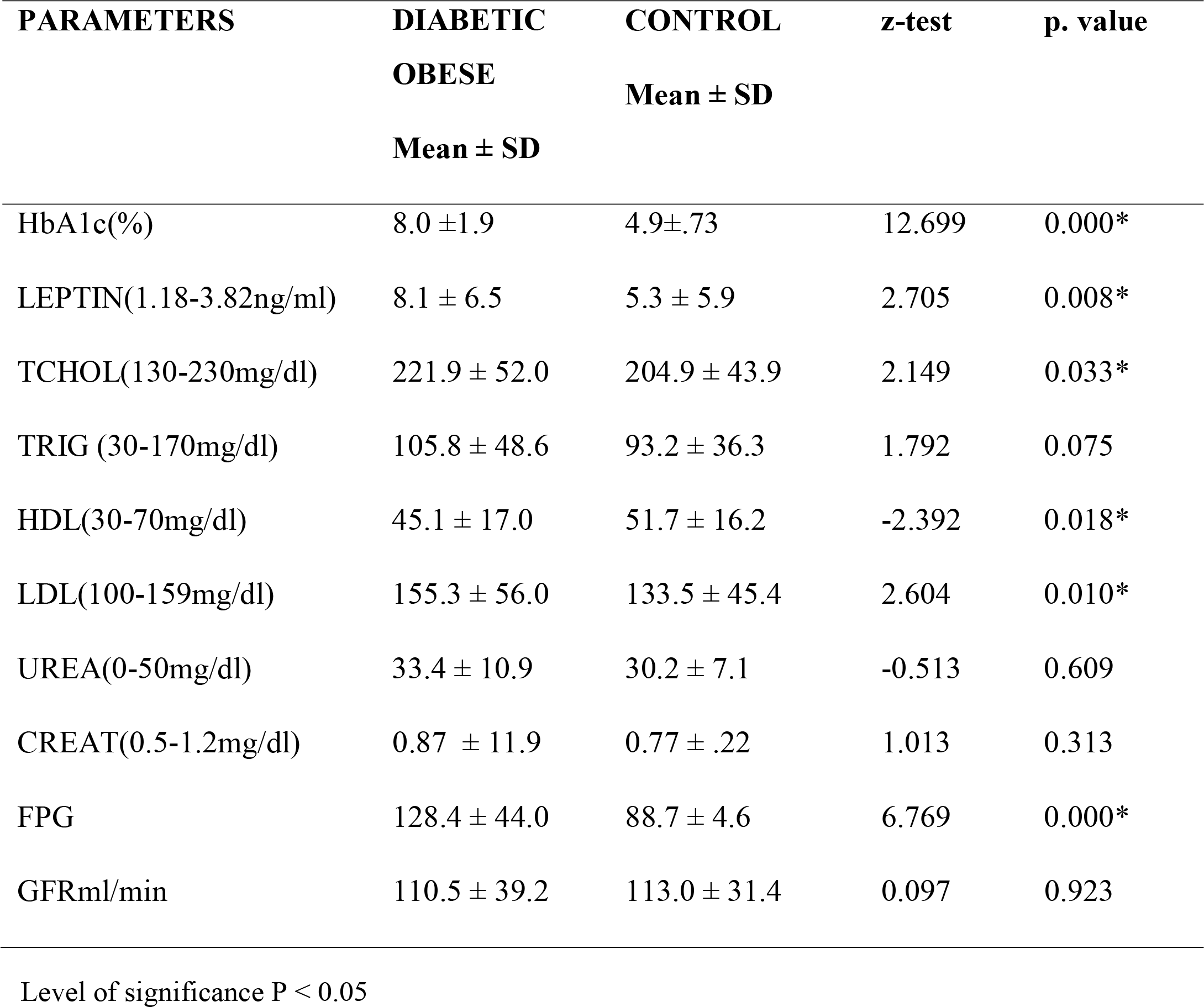
Biochemical parameters of DM obese and control subjects (Mean ± SD)

| **PARAMETERS** | **DIABETIC**<br>**OBESE**<br>**Mean ± SD** | **CONTROL**<br>**Mean ± SD** | **z-test** | **p. value** |
| --- | --- | --- | --- | --- |
| HbA1c(%) | 8.0 ± 1.9 | 4.9±.73 | 12.699 | 0.000* |
| LEPTIN(1.18-3.82ng/ml) | 8.1 ± 6.5 | 5.3 ± 5.9 | 2.705 | 0.008* |
| TCHOL(130-230mg/dl) | 221.9 ± 52.0 | 204.9 ± 43.9 | 2.149 | 0.033* |
| TRIG (30-170mg/dl) | 105.8 ± 48.6 | 93.2 ± 36.3 | 1.792 | 0.075 |
| HDL(30-70mg/dl) | 45.1 ± 17.0 | 51.7 ± 16.2 | -2.392 | 0.018* |
| LDL(100-159mg/dl) | 155.3 ± 56.0 | 133.5 ± 45.4 | 2.604 | 0.010* |
| UREA(0-50mg/dl) | 33.4 ± 10.9 | 30.2 ± 7.1 | -0.513 | 0.609 |
| CREAT(0.5-1.2mg/dl) | 0.87 ± 11.9 | 0.77 ± .22 | 1.013 | 0.313 |
| FPG | 128.4 ± 44.0 | 88.7 ± 4.6 | 6.769 | 0.000* |
| GFRml/min | 110.5 ± 39.2 | 113.0 ± 31.4 | 0.097 | 0.923 |

Level of significance $p < 0.05$

**Figure 4.1:**
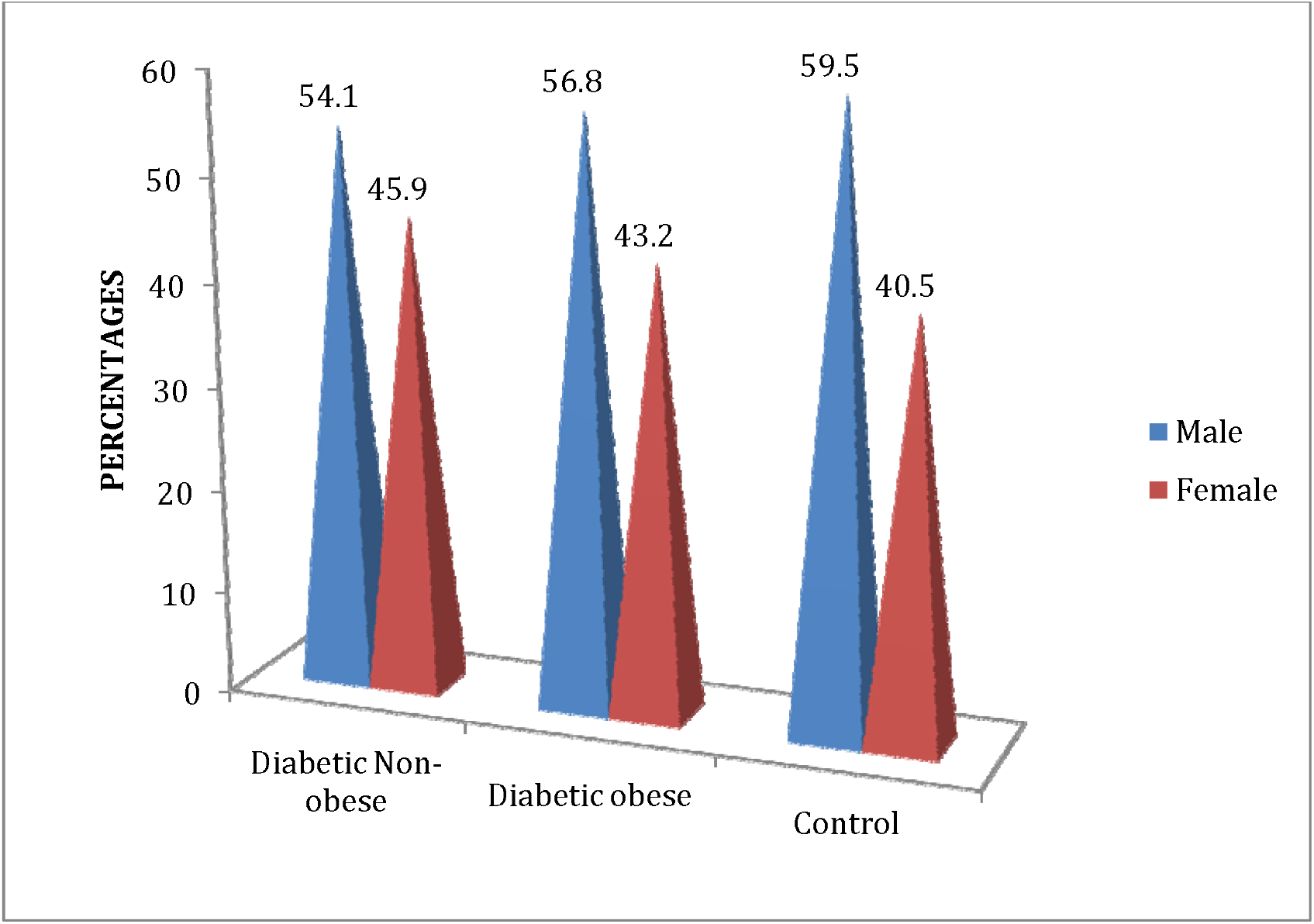
Percentage distribution of *gender groups* in test subjects and controls.

Table 2.4 shows the Pearson correlation coefficient between Leptin and Biophysical Parameter in the DM subjects (diabetic non-obese, diabetic obese) and controls. There was no significant association between leptin level against any of the biophysical parameters of the DM non-obese patients and controls. Leptin was significantly correlated with weight (r=0.194, p<0.05) in DM-obese patients. Leptin was significantly correlated with BMI (r=0.115, p<0.05) in DM-obese patients. Waist Hip Ratio was also found significantly correlated with Leptin (r =0.488, p<0.05) in DM-obese patients.

**Table 2.4.**
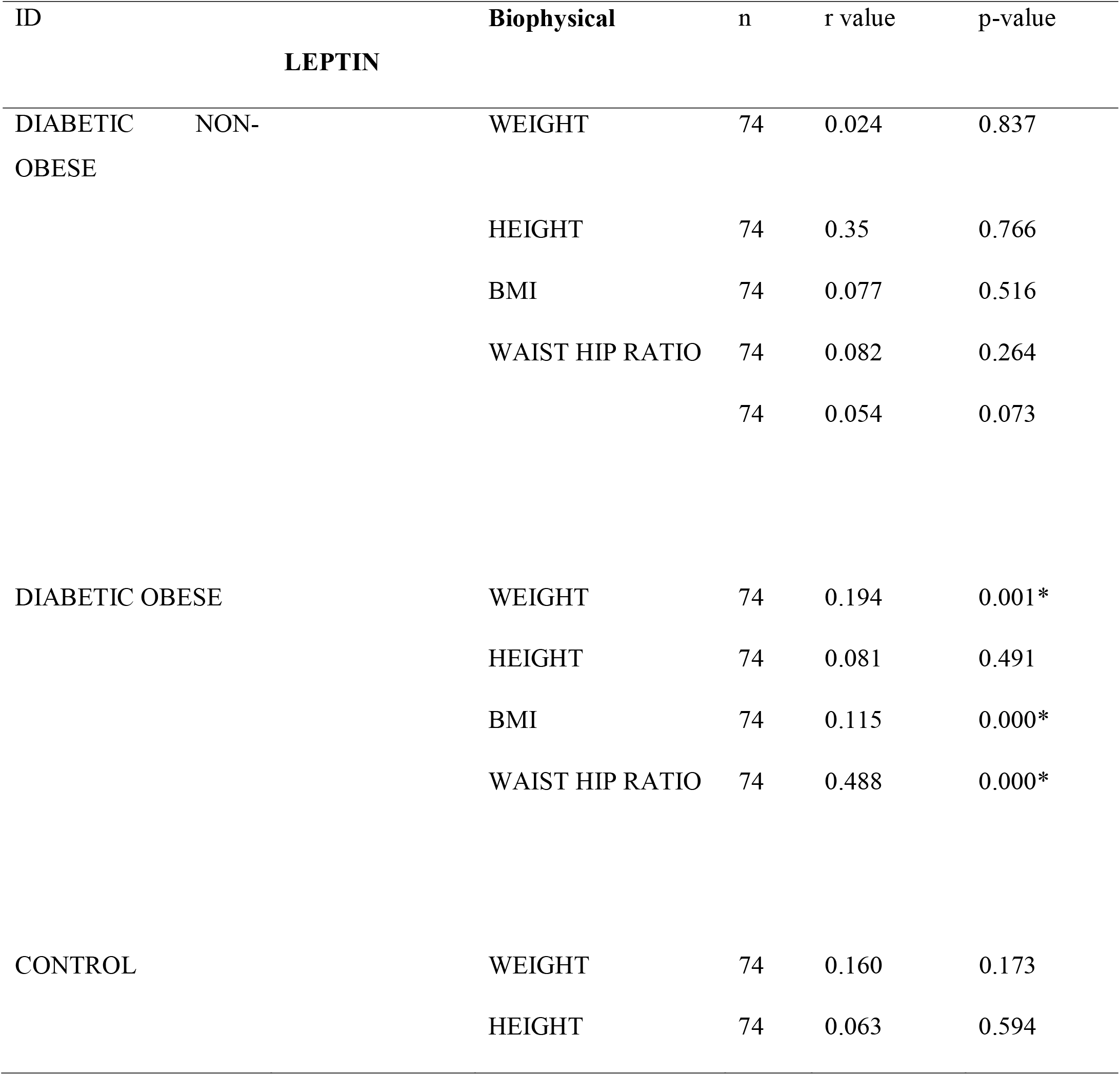
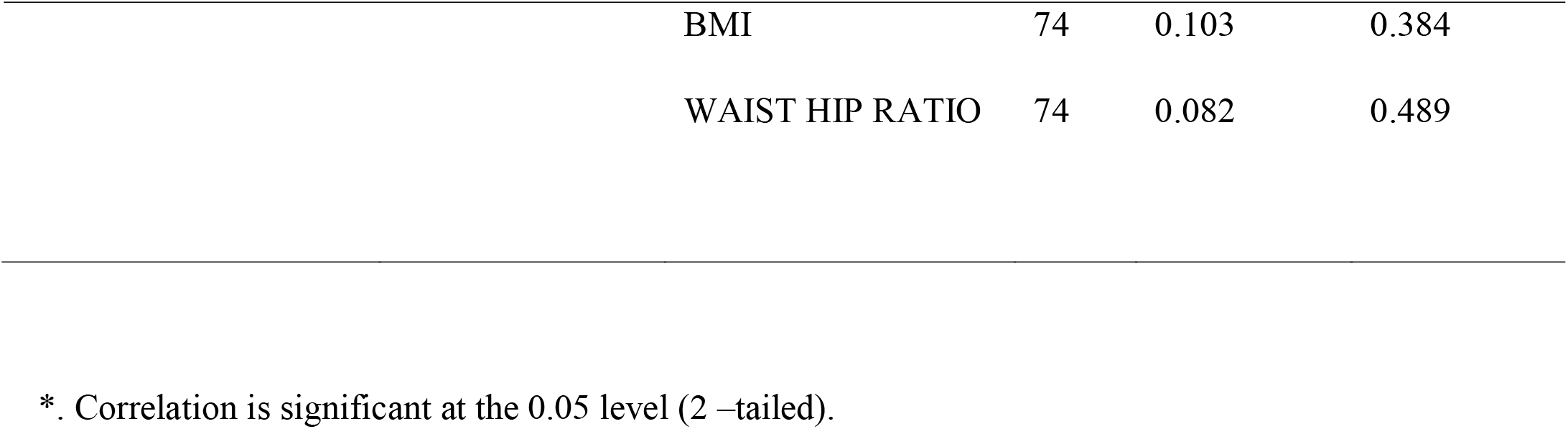
Correlations between Leptin and Biophysical Parameters in the DM Subjects (diabetic non-obese, diabetic obese) and Control.

Table 2.5 shows the Pearson correlation coefficient between Leptin and Lipid Profile in the DM subjects (diabetic non-obese, diabetic obese) and controls. There was no significant association between leptin level and the lipid profile of the DM non-obese patients and controls. Leptin was significantly correlated with Triglyceride (r=0.314, p<0.05) in DM-obese patients. Leptin was also significantly correlated with low-density lipoprotein(r=0.115, p<0.05) in DM-obese patients. Leptin was also found to have no significant correlation with total Cholesterol Cholesterol (r =0.118, p>0.05) and high-density lipoprotein (r =0.123, p>0.05) in DM-obese patients.

**Table 2.5.**
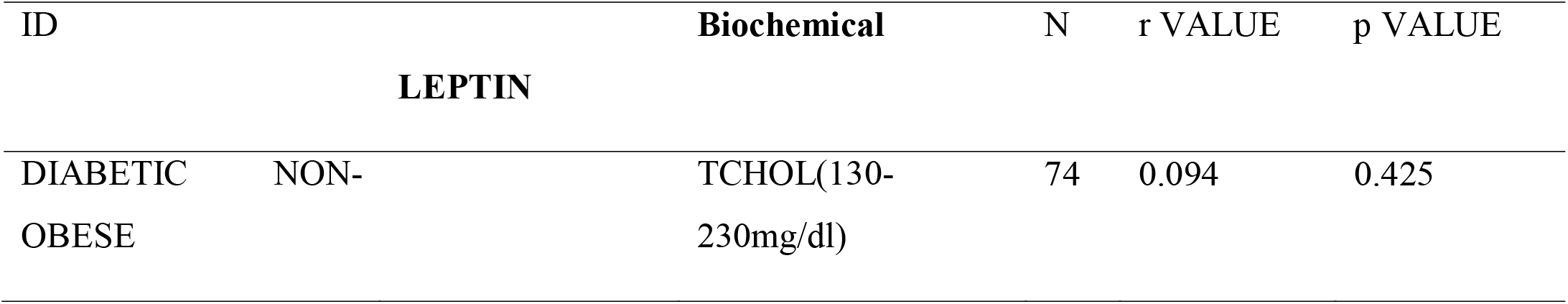
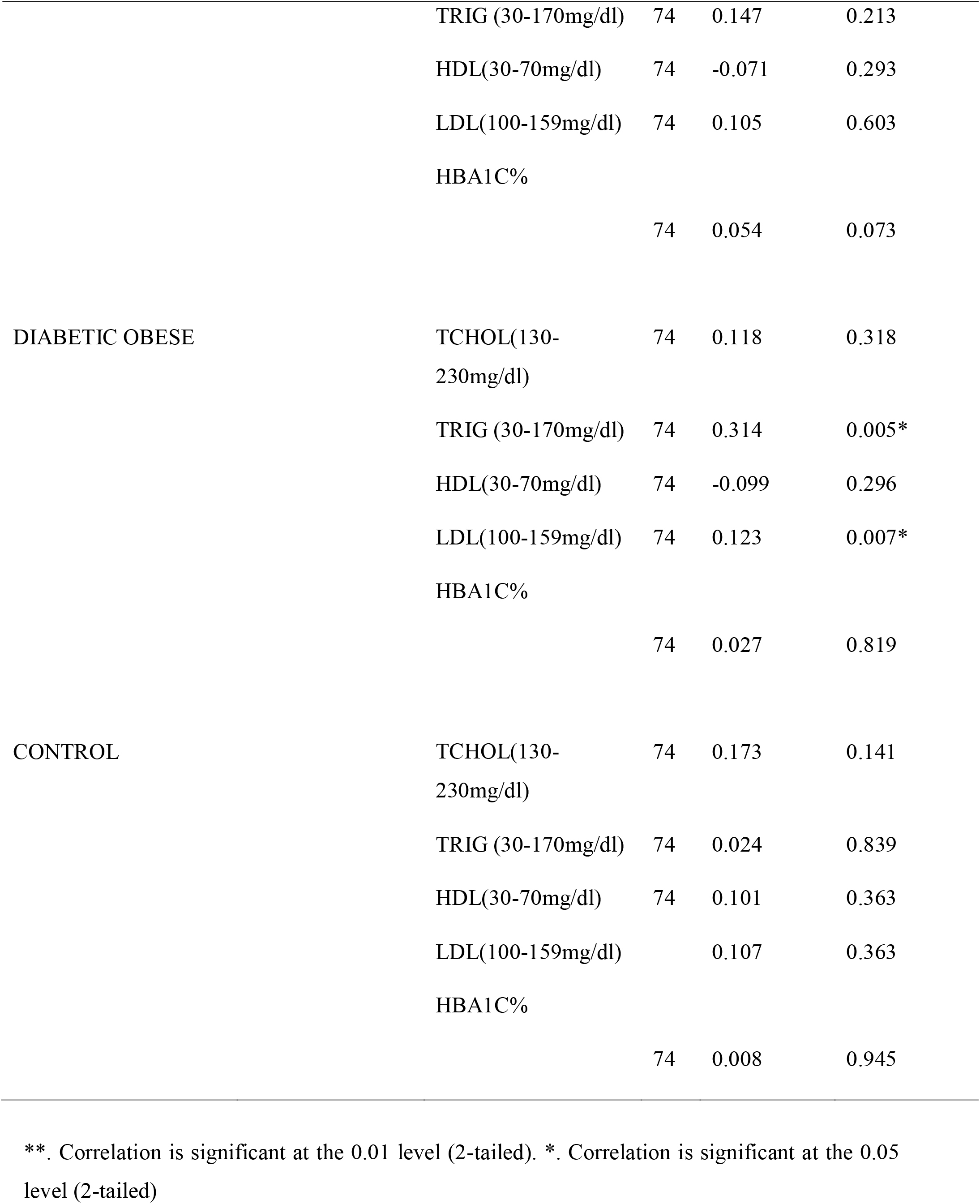
Correlations between Leptin and Lipid profile in the DM Subjects (diabetic non-obese, diabetic obese) and control.

**Table 2.6 shows the Pearson correlation coefficient between Leptin and Urea, Creatinine and eGFR of DM subjects (diabetic non-obese, diabetic obese) and controls.**

**Table 2.6:**
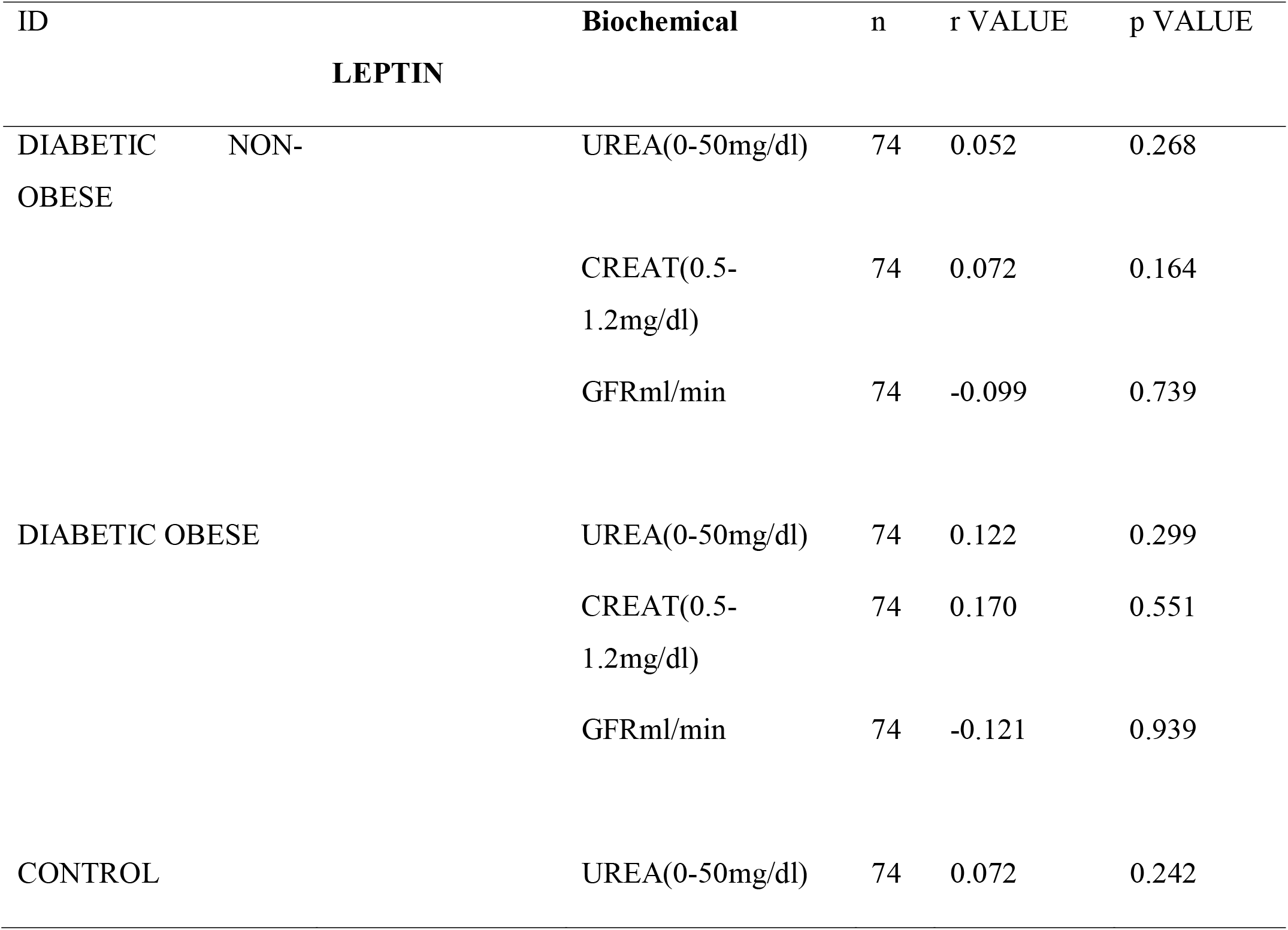
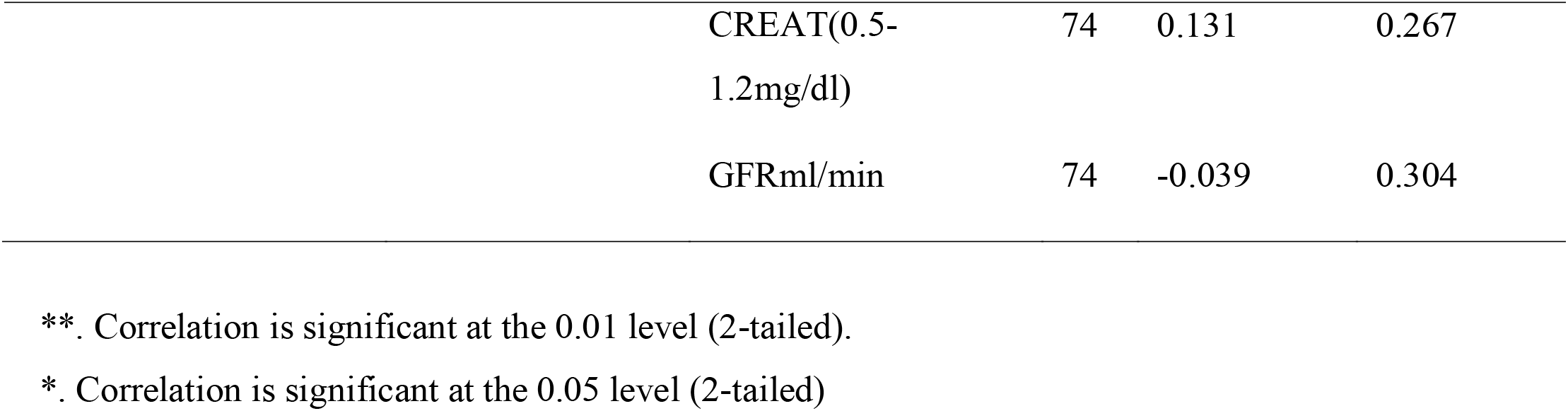
Correlations between Leptin and Urea, Creatinine and eGFR in the DM Subjects (diabetic non-obese, diabetic obese) and control.

| ID |  | Biochemical | n | r VALUE | p VALUE |
| --- | --- | --- | --- | --- | --- |
| **LEPTIN** |  |  |  |  |  |
| DIABETIC NON-OBESE |  | UREA(0-50mg/dl) | 74 | 0.052 | 0.268 |
|  |  | CREAT(0.5-1.2mg/dl) | 74 | 0.072 | 0.164 |
|  |  | GFRml/min | 74 | -0.099 | 0.739 |
| DIABETIC OBESE |  | UREA(0-50mg/dl) | 74 | 0.122 | 0.299 |
|  |  | CREAT(0.5-1.2mg/dl) | 74 | 0.170 | 0.551 |
|  |  | GFRml/min | 74 | -0.121 | 0.939 |
| CONTROL |  | UREA(0-50mg/dl) | 74 | 0.072 | 0.242 |

|  |  |  |  |
| --- | --- | --- | --- |
| CREAT(0.5-1.2mg/dl) | 74 | 0.131 | 0.267 |
| GFRml/min | 74 | -0.039 | 0.304 |

\*\*. Correlation is significant at the 0.01 level (2-tailed).
\*. Correlation is significant at the 0.05 level (2-tailed)

There was no significant association between Leptin level and the Urea, Creatinine and eGFR of the DM subjects and controls.

## DISCUSSION

Our study aimed to examine the profiles of blood leptin, lipids, HbA1C, and renal function in type 2 diabetes mellitus patients, with a particular emphasis on differences between non-obese and obese patients. To achieve this objective, a sample of 222 individuals was recruited, comprising 74 individuals diagnosed with type 2 diabetes mellitus but not classified as obese, 74 individuals classified as obese, and 74 healthy individuals serving as control subjects.

The results of our study provide valuable insights into the complex relationship between obesity and type 2 diabetes. Our findings demonstrate the need for comprehensive evaluations of these metabolic and biochemical markers in type 2 diabetes patients, particularly with respect to differences between non-obese and obese individuals. These insights are crucial for the effective clinical management of type 2 diabetes, a debilitating and increasingly prevalent chronic condition.

Based on the results of our study, a significant association has been established between obesity and type 2 diabetes in the studied population. Our findings, obtained through extensive data analysis and statistical evaluation, indicate that higher body mass index and waist-hip ratio are significantly associated with an increased risk of developing type 2 diabetes. This conclusion was drawn by comparing the differences between diabetic obese patients, diabetic non-obese patients, and a control group. These results have important implications for the early identification and management of type 2 diabetes and highlight the need for further research to understand better the underlying mechanisms linking obesity and type 2 diabetes.

Our study’s results align with a considerable body of evidence that has established a strong association between obesity and type 2 diabetes. For instance, a systematic review and meta-analysis by Galicia-Garcia et al., 2020, found that an elevated body mass index was significantly associated with an increased risk of type 2 diabetes (1). Similarly, a study by Huang et al., 2015, indicated that obesity, as evidenced by a high waist-hip ratio, was correlated with a heightened risk of type 2 diabetes. While obesity is a known risk factor for type 2 diabetes, several other factors, such as genetic predisposition, lifestyle habits, and other metabolic and hormonal imbalances, may also contribute to the development of the disease. Additionally, the precise mechanisms by which obesity contributes to the onset of type 2 diabetes remain to be fully understood and require further investigation.

In light of our study results, it is imperative to continue exploring the intricate relationship between demographic, anthropometric, metabolic, and hormonal factors in the aetiology and progression of type 2 diabetes, particularly in populations with a heightened risk of obesity. Further research is essential to deepen our understanding of the underlying mechanisms of type 2 diabetes and to design and implement effective strategies for its prevention and management.

Given the increasing prevalence of both obesity and type 2 diabetes, addressing this pressing public health challenge is of utmost importance. By conducting robust and comprehensive research, we can understand the interplay between these factors and develop targeted and effective interventions to reduce the burden of type 2 diabetes in affected populations.

Furthermore, our results showed that obesity could have a negative impact on lipid profile and glycemic control in these patients. Specifically, we found that while triglyceride levels did not differ significantly between the two groups, the obese group had significantly higher levels of HbA1c, Leptin, total CholesterolCholesterol, HDL, and LDL compared to the non-obese group. This indicates that obesity may contribute to the development of metabolic abnormalities in type 2 diabetes mellitus.

These findings are consistent with previous studies showing a positive correlation between obesity and HbA1c levels in type 2 diabetes mellitus patients (3, 4, 5). Similarly, research has reported elevated levels of total CholesterolCholesterol and LDL in obese patients with type 2 diabetes mellitus (4). The higher levels of Leptin in the obese group in our study also agree with prior studies that have established a connection between obesity and increased leptin levels.

In addition, our results indicated that obese individuals with type 2 diabetes exhibited elevated HbA1c, Leptin, Total Cholesterol, HDL, LDL, and fasting plasma glucose compared to non-obese individuals with the same condition. This difference was statistically significant (p < 0.05). The findings of our study concur with previous studies that have reported the negative impact of obesity on glycemic control and lipid profile in type 2 diabetes patients (6, 7). A study by Sherwani et al., 2016 reported that obesity was associated with higher levels of HbA1c and fasting plasma glucose in type 2 diabetes patients, which supports our findings (8). Another study by Hussain et al., 2019 found that obesity was linked to elevated levels of total CholesterolCholesterol, LDL, and triglycerides, consistent with our results regarding the impact of obesity on lipid profile in type 2 diabetes patients (9). However, in contrast to our findings, Amidu et al., 2019 reported no significant difference in HbA1c, triglycerides, and fasting plasma glucose levels between obese and non-obese type 2 diabetes patients (10). This discrepancy may be due to differences in study populations, sample sizes, or methodologies used in the studies.

The results of our recent study shed light on the differential relationship between leptin levels and lipid profiles in diabetic patients based on their obesity status. Our findings indicate that in diabetic non-obese patients and controls, there was no significant correlation between leptin and lipid profile parameters. However, a different picture emerged in diabetic obese patients, where a significant correlation was observed between leptin and triglyceride levels (r=0.314, p<0.05) as well as between leptin and low-density lipoprotein levels (r=0.115, p<0.05). There was no significant association between Leptin and total CholesterolCholesterol (r=0.118, p>0.05) or high-density lipoprotein (r=0.123, p>0.05) in diabetic obese patients. These findings align with previous studies that reported a positive correlation between leptin levels and triglyceride levels in obese populations (11, 12). On the other hand, some studies have reported a negative correlation between leptin and high-density lipoprotein levels, highlighting the complex and conflicting relationship between leptin levels and lipid profiles (13, 14).

It is important to note that the conflicting results of previous studies may be due to differences in study populations, sample size, and other confounding factors. Further research is needed to clarify the relationship between leptin levels and lipid profiles in diabetic patients, particularly those with obesity. This will provide important insights into the mechanisms underlying this relationship and inform the development of effective strategies for managing lipid profiles in diabetic patients.

The results of our study provide insights into the relationship between leptin levels and biophysical parameters in type 2 diabetes mellitus patients based on obesity status. Our findings indicate that in diabetic non-obese patients and controls, there was no significant correlation between leptin levels and any of the biophysical parameters measured. However, in diabetic obese patients, our results reveal a significant correlation between leptin levels and weight (r=0.194, p<0.05) as well as between leptin levels and body mass index (r=0.115, p<0.05). Additionally, we found a significant correlation between the waist-hip ratio and leptin levels (r=0.488, p<0.05) in this group.

These findings are consistent with previous studies that reported a significant association between leptin levels and obesity-related parameters in individuals with type 2 diabetes mellitus (4, 5). For instance, a study reported a positive correlation between leptin levels and waist circumference in Korean individuals with type 2 diabetes (7). Similarly, a study found a significant positive correlation between leptin levels and body mass index in Saudi Arabian individuals with type 2 diabetes (9). These results suggest that obesity may play a role in the relationship between leptin levels and biophysical parameters in type 2 diabetes mellitus patients.

Our study’s results indicated no statistically significant difference in urea, creatinine, and estimated glomerular filtration rate (eGFR) between diabetic obese and non-diabetic obese individuals. Conversely, a significant difference was observed in these parameters between the control group and the diabetic group. These findings align with previous studies that have established diabetes as a risk factor for decreased kidney function, as evidenced by elevated levels of creatinine and urea and a decrease in eGFR. For instance, a study found that patients with type 2 diabetes exhibited significantly lower eGFR compared to healthy controls (10).

Similarly, a study reported that diabetes was linked with an increased risk of kidney dysfunction, indicated by elevated creatinine and urea levels (11). Our study results suggest that obesity, in the absence of diabetes, may not be a risk factor for impaired kidney function in patients with type 2 diabetes. This is a noteworthy discovery, as previous research has provided conflicting results on the impact of obesity on kidney function in diabetes.

Further studies are needed to understand the underlying mechanisms of the observed differences in kidney function between diabetic and non-diabetic obese individuals and assess the generalizability of these results to other populations. Also, further research is needed to clarify the mechanisms underlying this relationship and inform the development of effective strategies for managing type 2 diabetes mellitus in obese individuals. However, further research is needed to confirm these results and to determine the underlying mechanisms responsible for the observed relationships between Leptin, obesity, and biophysical parameters in type 2 diabetes mellitus patients. This could include exploring the role of insulin resistance and inflammation in mediating these relationships and evaluating the potential impact of other factors, such as diet and physical activity, on these relationships.

Strengths of this study include a well-defined patient population, standardised measures to assess lipid profile and glycemic control, and a large sample size that increases the statistical power of the results. However, there are also limitations that must be considered. First, the study was cross-sectional, so causality cannot be established. Second, the sample was drawn from a single centre, so the generalizability of the results to other populations may be limited. Third, dietary and lifestyle factors that may impact lipid profile and glycemic control were not systematically assessed, and their potential impact on the results cannot be ruled out. The results should be interpreted in light of the study’s limitations, and future research is needed to investigate the underlying mechanisms further and determine the generalizability of the results to other populations.

## CONCLUSIONS

The conclusions of our study add to the growing body of evidence that underscores the importance of weight management as a key component in the management of type 2 diabetes. Our results indicate that obesity has a negative impact on lipid profile and glycemic control in type 2 diabetes patients and highlights the significance of considering the effect of obesity on the relationship between leptin and lipid profile in diabetic patients. These findings suggest that targeting obesity and leptin levels may be an effective strategy for improving lipid profile and glycemic control in type 2 diabetes. Further research is needed to confirm these findings and explore the underlying mechanisms contributing to these differences.

## Data Availability

All data produced in the present work are contained in the manuscript

## List of Abbreviations

BMI: Body Mass Index
WHR: Waist hip ratio
S.D: standard deviation
TCHOL: Total CholesterolCholesterol
TRIG: Triglyceride
HDL: high-density lipoprotein
LDL: low-density lipoprotein
HBA1C: Glycated haemoglobin
FPG: Fasting Plasma Glucose
eGFR: Estimated Glomerular Filtration rate
CREAT: Creatinine

## Acknowledgement

None

## Availability of Data

The dataset is available upon request from the authors

